# Exploring Burn First Aid Knowledge and Water Lavage Practices in Uganda: A Cross-Sectional Study

**DOI:** 10.1101/2023.08.10.23293067

**Authors:** Brian Kasagga, Joseph Baruch Baluku, Felix Bongomin, Derrick Kasozi, Eria Muwanguzi, Mercy Namazzi, Yusuf Sadiq, Rose Alenyo, Edris Wamala Kalanzi, Darius Balumuka, Alex Emmanuel Elobu

## Abstract

**Background:** Low- and middle-income countries experience higher burn-related morbidity and mortality compared to high-income countries. Prehospital Burn First Aid (BFA) has been proven effective in reducing such outcomes. This study aimed to assess BFA knowledge and water lavage practices and their associated factors among burn victims and the general population at a tertiary health facility in Uganda.

**Methods:** A cross-sectional study was conducted at the Burns Unit of Kiruddu National Referral Hospital in Kampala between April and November 2022. Participants included burn patients, caregivers, and visitors. Data on BFA knowledge and practices were collected using an interviewer-administered questionnaire. BFA knowledge was evaluated using 13 questions, with ≥80% score considered adequate. Logistic regression was used to analyze associations.

**Results:** The study had 404 participants, comprising 68(16.8%) burn victims, 161(39.9%) primary caregivers, and 175(43.3%) visitors. Among all participants, 339(83.9%) had never received BFA information, and 392(97.0%) had no first aid training. Mean BFA knowledge score was 56% (SD 13.9), with only 5.4% demonstrating adequate knowledge. Only 26(27.7%) of current and former burn victims used water lavage as BFA. No statistically significant associations were found between BFA knowledge, water lavage usage, and demographic variables on univariate and multivariate analyses.

**Conclusion:** This study highlights inadequate BFA knowledge and practices among participants. Addressing these deficiencies through community-based initiatives is crucial to improving burn care in Uganda.

## INTRODUCTION

Flames, hot liquids, hot gases, hot surfaces, cold, corrosive chemicals, electricity, lightning, and radiation can all result in burn injuries.^1^The severity of damage caused by burns is determined by the energy of the causative agent and the duration of exposure. The skin is the most commonly affected tissue during burns and accounts for the majority of damage. However, in some cases, electricity can cause more extensive tissue damage than what is visible on the skin. Internal damage to the airways can occur from inhalation of smoke or hot gases. Systemic derangements are frequently observed in cases of larger or deeper burns with a total burn surface area (TBSA) exceeding 30%.^1,2^

Burn injuries are common worldwide, with over 180,000 annual deaths. Most fatalities occur in low- and middle-income countries (LMICs), where data on burns is scarce, suggesting the actual number may be higher.^3^ In sub-Saharan Africa, scalds are the most common cause of burns, accounting for 59% of burns, while flame burns account for 33%.^4^ In Uganda, scalds are most common in male children under the age of five, accounting for approximately 11% of unintentional injuries in this age group.^5^ Acid casualties, on the other hand, result from assault and occur in older individuals with a median age of 33.^6^

Morbidity and mortality from burns are particularly higher in LMICs compared to high-income countries(HICs). This is because HICs have implemented evidence-based remedies such as smoke alarms, water heater temperature control, early response, flame retardant children’s sleepwear, and effective burn first aid (BFA) and treatment practices ^7^ A global burden of disease analytical study by Yakupu et al. found that the years of life lost to premature mortality (YLL) and years of healthy life lost due to disability (YLD) due to burn injuries were 67% and 33%, respectively.^8^ Burn victims often suffer from lasting disabilities, disfigurement, and psychiatric disorders, along with social stigma and rejection. Long-term consequences can include an increased risk of cancer and emotional trauma, especially in children, who may be negatively impacted by scarring, hair loss, and restricted mobility. These physical and psychological effects can cause difficulties in social interactions, particularly for children who may miss out on social play.^4^

Burn injuries impose a significant economic burden on LMICs, with treatment costs higher than those for diseases like tuberculosis and HIV.^9,10^ In sub-Saharan Africa alone, burn-related losses exceeded $1 billion in 2019, and South Africa spent $26 million treating burns caused by kerosene and charcoal stoves.^11^ However, funding for burns is often inadequate, as illustrated by the Initiative for Social Economic Rights (ISER) report indicating that Uganda’s burn budget was among the unfunded priorities for 2020-21. External actors, such as non-governmental organizations and private actors, typically provide most of the funding for burns.^12,13^ Burns therefore place an additional burden on LMIC governments and healthcare systems, which are already struggling with insufficient funding. The socio-economic impact of burns is compounded by factors such as lost wages, prolonged care for deformities, and emotional trauma.^3^

Burns consume many surgical resources, often requiring repeated surgeries. With the limited surgery resources in sub-Saharan countries, the safe bet is to prevent burns in the first place, improve preparedness and reduce the extent of injury through proper prehospital care (burn first aid). Prehospital Burn First Aid (BFA) has been shown to reduce morbidity and mortality, as well as associated healthcare costs, by reducing tissue damage, the need for surgery, and the overall outcomes.^14-16^ In Uganda, as in most other sub-Saharan countries, burn victims use alternative practices such as herbs due to a lack of access to medical care. ^17^Even so, poor first aid practices, such as mud and cow dung, on the contrary, can lead to a worsening of the burn injury with adverse consequences. ^17-21^

To the best of our knowledge, studies evaluating BFA knowledge and practices in burn victims and the general Ugandan population are extremely scarce, with only a few retrospective chart-review studies. ^20,22^ Therefore, we aimed to examine BFA knowledge, BFA practices and their associated factors. These results will help us to design educational strategies for BFA.

## METHODS

### Design

A cross-sectional descriptive study using quantitative techniques was conducted between July 2022 and September 2022 in the Emergency, Inpatient, and Outpatient Departments (EPD, IPD, and OPD) of the Burns Unit, Kiruddu National Referral Hospital (KNRH).

### Study setting

The study was conducted at KNRH, one of Uganda’s largest hospitals. It offers burns, plastic surgery, radiology, internal medicine, and palliative care services. It is also a Plastic Surgery Teaching Hospital for the College of Surgeons of East and Central Africa (COSECSA) and the Department of Surgery, Makerere University College of Health Sciences. The hospital is located in Makindye Division, Kampala, approximately 13 km by road southeast of Mulago National Referral Hospital. It is Uganda’s highest tertiary hospital in the treatment of burns and has a monthly patient volume of approximately 60 patients.

### Study population and sample size

The sample size was calculated using the formula; n = (z/p)^2^π(1-π); we assumed that the population proportion with adequate BFA is about 50%, a 95% confidence interval. Therefore, n = (1.96/0.05)20.5(1-0.5) = 384. Having considered a possible 5% non-response rate, 404 people were enrolled by convenience sampling. All burn patients, their caretakers, and hospital visitors aged 12 years and above in the emergency and burn wards of KNRH were asked to participate in the study voluntarily.

### Data collection, sampling procedures and study measurements

Data was collected using a standardized questionnaire administered by the interviewers through a face-to-face interview. Through consecutive sampling, all ED, OPD, and IPD burn subjects who met the eligibility criteria were recruited into the study after informed consent. Their BFA knowledge was assessed using the questionnaire. The independent variables in this study included demographics (sex, age, education level, level of education, etc.), receipt of prior BFA information, prior burn experience, first aid training, and prior experience in providing care for burn victims. The dependent variables were BFA knowledge and BFA practices. The participants’ level of knowledge was deemed adequate if they scored 80% or higher on a 13-question questionnaire (Appendix 1) about appropriate first aid for various burn scenarios, including scalds, electricity, and acid. Each correct answer was worth 1 point and incorrect answers were scored 0. The total score was 13, and a calculated score of 80 percent or higher was considered adequate knowledge.

Appropriate practices were assessed through a question about practices following a burn, with a correct response being water usage and an incorrect response being non-usage of water. The questionnaire can be found in Appendix 1 and was developed based on a literature review and reviewed by plastic surgeons at KNRH who are experts in the field.

The data collection tool was pre-tested on ten people.

### Data management and analysis

After data collection, the data was entered into ready-made Google forms. The extracted data were cleaned and sorted using Microsoft Excel. The data were then analyzed using SPSS software for Windows version 26 (IBM, New York, NY). The data were summarized with descriptive statistics; Mean and SD for approximately normally distributed data and median and interquartile ranges for non-normally distributed data. Categorical data were presented as frequency and percentage. We used logistic regression analysis to determine factors associated with low BFA knowledge levels. P values < 0.05 were considered statistically significant.

### Participant consent and ethics approval

The study was approved by the Mulago Hospital Research and Ethics Committee (REC No; MHREC2226). The study was conducted following the Declaration of Helsinki guidelines. Written informed consent was obtained from participants aged 18 years and above, while assent was obtained from participants below 18 years in addition to parental/guardian consent.

## RESULTS

### Characteristics of study participants

A total of 404 participants responded to the survey, of which 68(16.8%) were current burn victims, 161(39.9%) were current primary caregivers of burn victims, and 175 (43.3%) were visitors. 186 (46%) participants had previously experienced a burn, while 54% of respondents had previously cared for burn victims. The larger percentage of respondents were women (55.9%, n=226). 46.0%(n=186) of those surveyed lived in Central Uganda. Regarding educational status, the largest share of respondents had completed high school (39.9%, n=161). The majority of respondents previously had neither received any BFA information (83.9%, n=339) nor participated in a first aid training course with burns components (97.0%,n=392). Table 1 summarises the characteristics of the participants

**Table 1.0:** Demographics of participants.

| **Characteristics** | **Frequency** | **Percentage** |
| --- | --- | --- |
| **Age in years** |  |  |
| 12-18 | 60 | 14.9 |
| 19-35 | 231 | 57.2 |
| 36-59 | 106 | 26.2 |
| Above 60 | 7 | 1.7 |
| **Gender** |  |  |
| Male | 178 | 44.1 |
| Female | 226 | 55.9 |
| **Nationality** |  |  |
| Ugandan | 402 | 99.5 |
| Non-Ugandan | 2 | 0.5 |
| **Region of residence in Uganda** |  |  |
| Central | 261 | 64.6 |
| Northern | 31 | 7.7 |
| Eastern | 47 | 11.6 |
| Western | 65 | 16.1 |
| **Area of residence** |  |  |
| Rural | 154 | 38.1 |
| Urban | 250 | 61.9 |
| **Highest education** |  |  |
| Less than primary | 46 | 11.4 |
| Primary | 143 | 35.4 |
| High school | 161 | 39.9 |
| College | 31 | 7.7 |
| University | 21 | 5.2 |
| Masters | 2 | 0.5 |
| **Current job** |  |  |
| Private work | 126 | 31.2 |
| Employed | 91 | 22.5 |
| Doesn't work | 187 | 46.3 |
| **Received any BFA information before?** |  |  |
| Yes | 65 | 16.1 |
| No | 339 | 83.9 |
| **Participated in BFA training before?** |  |  |
| Yes | 12 | 3.0 |
| No | 392 | 97.0 |
| **Experienced a burn injury before?** |  |  |
| Yes | 186 | 46.0 |
| No | 218 | 54.0 |
| **Have you ever cared for a person with a burn?** |  |  |
| Yes | 218 | 54.0 |
| No | 186 | 46.0 |

### Burn First Aid Knowledge

The mean BFA knowledge score was 56 % (SD: 13.9). Only 5.4 % (n= 22) of those surveyed scored ≥80% and were considered to have adequate knowledge of burn first aid. Only 31.7% (31.7%) indicated water lavage as a method of BFA and 43.6%(n=176) answered that they would use antibiotics before coming to hospital. However, only 10% knew the “drop and roll” technique for clothing fires. Regarding knowledge on using water for acid attacks, only 40 (9.9%) answered correctly as indicated in Table 2.0 below.

**Table 2.0:** Percentage of correct answers for the burn knowledge-related questions.

| **Burn knowledge Question** | **Expected correct answer** | **Number of correct responses** | **Percentage** |
| --- | --- | --- | --- |
| Burns can cause permanent injuries | Agree | 387 | 95.8 |
| Burns can cause mental disorders | Agree | 324 | 80.2 |
| In case of burn injury covering the burned area before heading to hospital can decrease infection risk | Agree | 254 | 62.9 |
| In case of burn injury, Picking blisters is an incorrect action | Agree | 224 | 55.4 |
| In case of burn injury Applying first aid at home leads to a better outcome | Agree | 373 | 92.3 |
| It is beneficial to use antibiotics for management | Disagree | 228 | 56.4 |
| Incase of electricity burn injury I should not touch the person if he/she is still in electrical current | Agree | 393 | 97.3 |
| Incase of electricity burn injury, the first action is to turn off electricity if possible | Agree | 392 | 97.0 |
| 2-year-old boy pulls a kettle of boiling water onto himself | Remove all clothes, and cool with running water for 20 minutes | 97 | 24.0 |
| 10 year old, wanders barefooted into a burning bush as family prepares garden. | Immediately access cool running water and apply for 20 minutes | 95 | 23.5 |
| A 25-year-old man is at a party, he is standing close to a candle and his shirt catches fire | Leave all clothing intact and instruct him to drop to the ground and roll over | 47 | 11.6 |
| 30 year old woman cleaning her swimming pool, acid splashes on her face. What do you do? | Remove clothing and cool in water, shower for 20 min | 40 | 9.9 |
| First aid for a scald | Water lavage | 128 | 31.7 |

### Factors associated with BFA knowledge

In total, 382 respondents (94.6%) were found to possess inadequate BFA knowledge. Of these, 370(95.9%) of them had never received first aid training. The greater percentages of these individuals were from the central region (77.3%, n=17), were female (68.2%, n=15), and were aged between 19 to 35 years (59.1%, n=13). Additionally, the majority were visitors (44.0%, n=168), and had never experienced burns (53.1%, n=203). However, none of these relationships were significant in both bivariate and multivariable analyses, as demonstrated in Table 3.0.

**Table 3.0:** The association between BFA knowledge and demographic variables.

| Demographics | Total score<br>Mean(SD) | BFA knowledge,<br>N(%) |  | Logistic<br>Regression |  |  |  |
| --- | --- | --- | --- | --- | --- | --- | --- |
|  |  | Inadequate | Adequate | cOR | P | aOR (95%<br>CI) | P |
| **Overall** | 7.4(1.8) | 382(94.6) | 22(5.4) | NA | - | NA | - |
| **Type of respondent** |  |  |  |  |  |  |  |
| Burn victim | 7.0(1.6) | 66(17.3) | 2(9.1) | 1 |  | 1 |  |
| Care-taker | 7.1(1.5) | 148(38.7) | 13(59.1) | 2.90(0.64-13.21) | 0.169 | 3.25(0.63-16.91) | 0.161 |
| Visitors | 7.1(1.7) | 168(44.0) | 7(31.8) | 1.38(0.278-6.79) | 0.696 | 1.41(0.23-8.05) | 0.696 |
| **Sex** |  |  |  |  |  |  |  |
| Female | 7.1(1.6) | 211(51.2) | 15(68.2) | 1.737(0.69-4.36) | 0.25 | 0.57(0.21-1.52) | 0.26 |
| Male | 7.1(1.6) | 171(44.8) | 7(31.8) | 1 |  | 1 |  |
| **Age** |  |  |  |  |  |  |  |
| 12 to 18 | 6.7(1.9) | 57(14.9) | 3(13.6) | 1 |  | 1 |  |
| 19 to 35 | 7.1(1.6) | 218(51.1) | 13(59.1) | 1.133(0.31-4.11) | 0.85 | 0.79(0.18-3.51) | 0.76 |
| 36 to 59 | 7.2(1.5) | 100(26.2) | 6(27.3) | 1.14(0.275-4.73) | 0.86 | 0.78(0.15-3.95) | 0.76 |
| ≥60 | 6.7(0.8) | 7(1.8) | 0(0) | NA | NA | NA | NA |
| **Region** |  |  |  |  |  |  |  |
| Central | 7.3(1.6) | 244(63.9) | 17(77.3) | 1 |  | 1 |  |
| Northern | 6.6(1.4) | 30(7.9) | 1(4.5) | 0.48(0.06-3.72) | 0.48 | 0.457(0.05-4.17) | 0.49 |
| Eastern | 6.5(1.7) | 44(11.5) | 3(13.6) | 0.98(0.28-3.48) | 0.97 | 1.13(0.24-5.26) | 0.88 |
| Western | 6.7(1.5) | 64(16.8) | 1(4.5) | 0.22(0.029-1.72) | 0.15 | 0.155(0.02-1.35) | 0.09 |
| **Occupation** |  |  |  |  |  |  |  |
| Unemployed | 6.7(1.6) | 178(46.6) | 9(40.9) | 1 |  | 1 |  |
| Private work | 7.1(1.6) | 117(30.6) | 9(40.9) | 1.10(0.33-3.67) | 0.877 | 2.34(0.79-6.9) | 0.125 |
| Employed | 7.3(1.6) | 87(22.8) | 4(18.2) | 1.67(0.50-5.61) | 0.404 | 1.1(0.30-4.07) | 0.88 |
| **Level of education** |  |  |  |  |  |  |  |
| Primary | 6.7(1.4) | 177(46.3) | 12(54.5) | 1 |  | 1 |  |
| Secondary | 7.3(1.7) | 184(48.2) | 8(36.4) | 0.64(0.26-1.61) | 0.34 | 0.61(0.23-1.66) | 0.33 |
| Tertiary | 7.7(1.6) | 21(5.5) | 2(9.1) | 1.4(0.29-6.71) | 0.67 | 5.04(0.72-35.2) | 0.10 |
| **Area of residence** |  |  |  |  |  |  |  |
| Rural | 6.8(1.4) | 147(38.5) | 7(31.8) | 1 |  | 1 |  |
| Urban | 7.3(1.7) | 235(61.5) | 15(68.2) | 1.34(0.53-3.37) | 0.533 | 1.29(0.40-4.13) | 0.67 |
| **Experienced Burn Before** |  |  |  |  |  |  |  |
| Yes | 7.2(1.6) | 179(46.9) | 7(31.8) | 0.53(0.21-1.33) |  | 0.53(0.21-1.33) | 0.18 |
| No | 6.9(1.6) | 203(53.1) | 15(68.2) | 1 | 0.18 |  |  |
| **Have you ever received BFA information?** |  |  |  |  |  |  |  |
| Yes | 7.7(1.8) | 63(16.5) | 2(9.1) | 0.506(0.12-2.22) | 0.34 |  |  |
| No | 6.9(1.5) | 319(83.5) | 20(90.9) |  |  |  |  |
| **Have you ever received BFA training?** |  |  |  |  |  |  |  |
| Yes | 7.6(1.4) | 12(3.1) | 0(0) | NA | NA | NA | NA |
| No | 7.0(1.6) | 370(96.9) | 22(100) |  |  |  |  |
| **Have you ever cared for a Burns victim?** |  |  |  |  |  |  |  |
| Yes | 7.3(1.5) | 179(46.9) | 7(31.8) | 0.53(0.21-1.33) |  |  |  |
| No | 6.9(1.6) | 203(53.1) | 15(68.2) | 1 | 0.18 |  |  |

Total score out of 13. Adequate score was ascertained with Blooms cut off greater than 80 percent.

### Factors assosciated with water lavage usage following burns among current or former burn victims

Only 26(27.7%) of current and former burn victims used water lavage as BFA. Table 4 shows the outcomes of both bivariate and multivariate analyses.. Overall, the relationships between demographic variables and water lavage usage did not demonstrate significant associations on either bivariate or multivariate analyses. Rather than water, respondents used various substances as shown in figure 1.0

**Table 4.** Factors associated with water lavage usage in current and previous burn patients.

| Demographics | BFA water lavage practices |  | Logistic Regression |  |  |  |
| --- | --- | --- | --- | --- | --- | --- |
|  | Non-water lavage usage | Water lavage use. | cOR | P | aOR (95% CI) | P |
| **Overall** | 68(72.3) | 26(27.7) | NA |  | NA |  |
| **Sex** |  |  |  |  |  |  |
| Male | 35(51.5) | 11(42.3) | 1 |  |  |  |
| Female | 33(48.5) | 15(57.7) | 1.45(0.58-3.60) | 0.428 | 0.65(0.19-2.16) | 0.483 |
| **Age** |  |  |  |  |  |  |
| 12 to 18 | 17(25.0) | 7(26.9) | 1 |  | 1 |  |
| 19 to 35 | 34(50.0) | 16(61.5) | 1.14 (0.40-3.30) | 0.805 | 0.90(0.22-3.37) | 0.883 |
| 36 to 59 | 17(25.0) | 3(11.5) | 0.43(0.01-1.94) | 0.272 | 0.10(0.01-1.48) | 0.094 |
| ≥60 | 0(0) | 0(0) | NA | NA | NA |  |
| **Region** |  |  |  |  |  |  |
| Central | 46(67.6) | 23(88.5) | 1 |  | 1 |  |
| Northern | 3(4.4) | 1(3.8) | 0.67(0.07-6.77) | 0.732 | NA |  |
| Eastern | 8(11.8) | 0(0) | NA |  | NA |  |
| Western | 11(16.2) | 2(7.7) | 0.36(0.07-1.78) | 0.212 | 0.18(0.02-1.74) | 0.137 |
| **Occupation** |  |  |  |  |  |  |
| Unemployed | 32(47.1) | 12(46.2) | 1 |  | 1 |  |
| Private work | 24(35.3) | 4(15.4) | 0.45(0.15-1.31) | 0.143 | 0.35(0.07]-1.79) | 0.207 |
| Employed | 12(17.6) | 10(38.5) | 0.2(0.05-0.77) | 0.200 | 2.69(0.67-10.53) | 0.155 |
| **Level of education** |  |  |  |  |  |  |
| Primary | 38(55.9) | 11(42.3) | 1 |  | 1 |  |
| Secondary | 28(41.2) | 14(53.8) | 1.73(0.68-4.37) | 0.249 | 1.10(0.34-3.56) | 0.875 |
| Tertiary | 2(2.9) | 1(3.8) | 1.73(0.14-20.89) | 0.667 | 2.05(0.07-63.4) | 0.683 |
| **Area of residence** |  |  |  |  |  |  |
| Rural | 22(32.4) | 6(23.1) | 1 |  | 1 |  |
| Urban | 46(67.6) | 20(76.9) | 1.59(0.56-4.53) | 0.380 | 0.12(0.01-1.09) | 0.059 |
| **Experienced Burn Before** |  |  |  |  |  |  |
| No | 27(39.7) | 12(46.2) | 1 |  | 1 | 0.728 |
| Yes | 41(60.3) | 14(53.8) | 0.77(0.11-1.18) | 0.571 | 0.79(0.21-2.94) |  |
| **Have you ever received BFA training?** |  |  |  |  |  |  |
| No | 46(67.6) | 15(57.7) | 1 |  | NA | NA |
| Yes | 22(32.4) | 11(42.3) | 8.74(0.87-88.24) | 0.066 |  |  |
| **Have you ever received any BFA information?** |  |  |  |  |  |  |
| No | 60(88.2) | 21(80.8) | 1 | 0.571 | 1 | 0.819 |
| Yes | 8(11.8) | 5(19.2) | 0.77(0.31-1.91) |  | 0.78(0.09-6.85) |  |
| **Have you ever cared for a Burns victim?** |  |  |  |  |  |  |
| No | 67(98.5) | 23(88.5) | 1 |  | 1 | 0.182 |
| Yes | 1(1.5) | 3(11.5) | 1.53(0.61-3.88) | 0.367 | 2.67(0.63-11.3) |  |

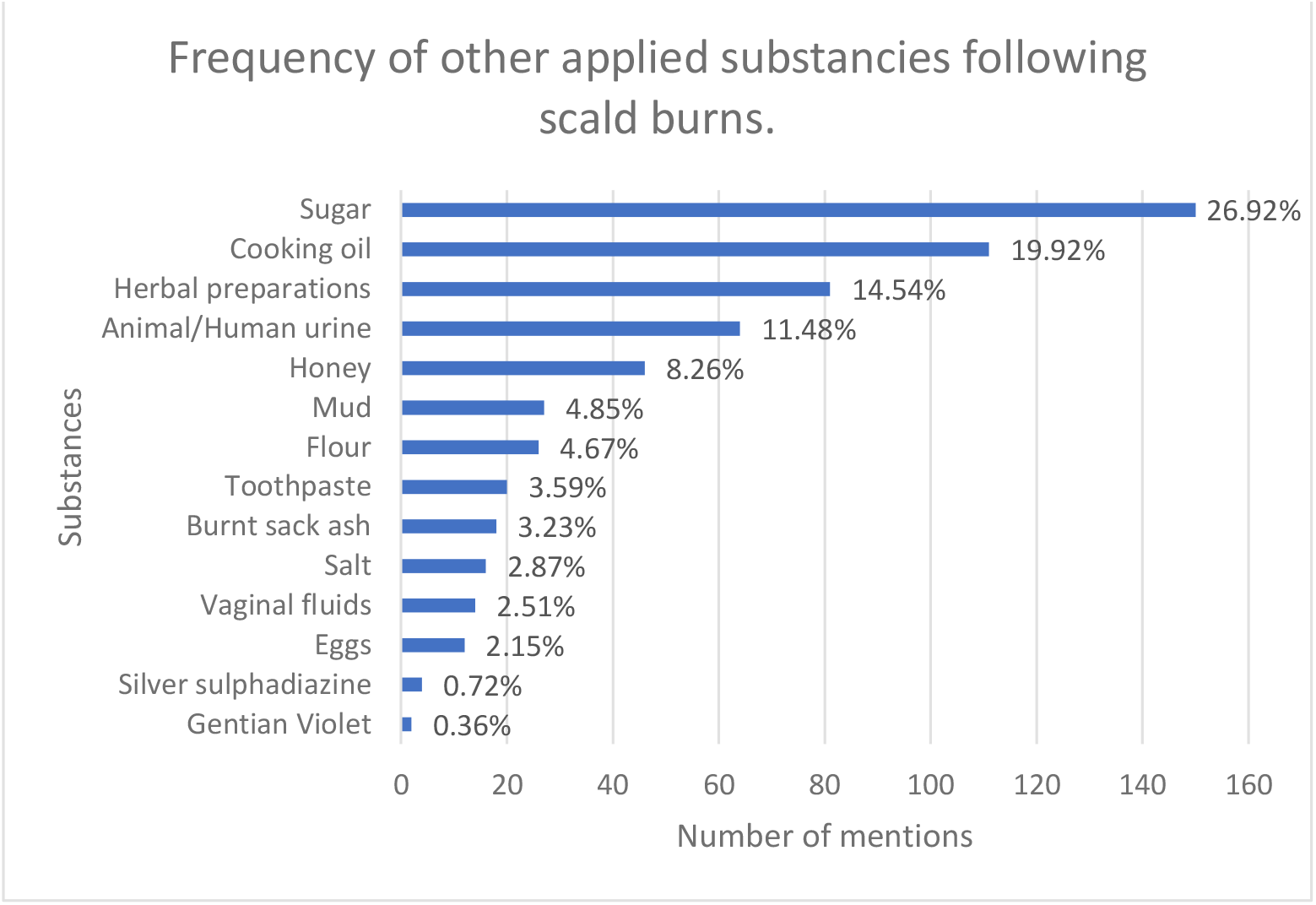

## DISCUSSION

This study aimed to assess BFA knowledge and water lavage practices and their associated factors. BFA can improve burn outcomes by reducing tissue damage, hospitalization time, and surgical interventions required.

General BFA knowledge was very low and only 5.4% (n=22) of participants had adequate BFA knowledge. This was consistent across all participants, regardless of whether they were currently a burn victim or not, with an average score of 56% on BFA knowledge questions This result (94.6%) is much higher compared with other studies done in Ethiopia, Indonesia, India where those with poor knowledge were 66.2, 66, and 60 percent, respectively^23-25^. However, our figure is higher because burn victims, caretakers, and visitors of burn patients in the hospital setting were all interviewed in this study, which increased the likelihood that people who participated in this study lacked knowledge to begin with. Furthermore, we utilized a Bloom’s cut-off score of greater than 80 percent to assess the adequacy of BFA knowledge, as opposed to the greater than 50 percent in the above studies.

Our study found that an overwhelming majority of participants (over 97%) were aware of the correct first aid steps to take during electric shock incidents, such as avoiding direct contact and promptly turning off the electricity. However, it was surprising to discover that only a small percentage (10%) were familiar with the “drop and roll” technique in case of clothing catching fire. This awareness level was almost significantly lower compared to studies conducted in Pakistan (20.5%)^26^, Bangladesh(80.7)^27^ and Vietnam (75%).^28^ This could be probably because of fewer education and safety campaigns regarding fires in Uganda, as indicated by a previous study where they found majority of schools were unprepared to deal with fires.^29^

However, concerning the use of antibiotics, almost half of the respondents (n=176, 43.6%) answered yes to utilizing antibiotics prior to seeking medical attention. This pattern resembles the practice of administering antibiotics without proper evaluation from healthcare professionals, which can be counterproductive and contribute to the development of antibiotic resistance.^30^ Lastly, the significance of water lavage in acid attack cases was recognized by a mere 10%. These findings shed light on the need for widespread education and awareness campaigns to address gaps in knowledge regarding essential first aid practices, particularly those pertaining to antibiotic use, fire safety, and acid attack response.

According to the published work, the current recommendations for first aid treatment of burn injuries advocate using cold water lavage (between 2 and 15°C) for 15-20 minutes. ^16^While over 92% of respondents recognized the benefits of applying first aid at home, the actual practices among burn victims and those with recent burn history were concerning. Only 22.1% (n=15) of admitted burn patients applied cold water lavage, which is lower compared to similar studies conducted in Ethiopia and Nigeria.^23,31^ The use of traditional mirrored patterns observed in the other developing countries, but these practices have no supporting evidence for their efficacy.^23,31,32^ It is most likely that these substances help to just soothe or numb the pain associated with the burns momentarily.

Furthermore, the utilization of these alternative burn first aid practices complicates wound evaluation and management. ^33,34^This is evident in a study conducted in Pakistan, where a substantial number of patients present with severely infected wounds that necessitate excision and skin grafting due to ineffective home treatments.^35^

## Strengths and Limitations

The study included a limited subgroup of only 68 burn victims, which constrained the exploration of practices specific to this group. We thus added 26 respondents who had experienced burns within the last 12 months. The overall respondent pool consisted of 404 participants, allowing for an assessment of burn first aid (BFA) knowledge among all respondents. It is important to note that this study may be susceptible to recall bias, and this factor should be considered when interpreting the results. We therefore tried to address this by assessing water lavage practices for those who got burn in the last 12 months. While the burn first aid questionnaire was developed based on a comprehensive literature review, it was not a validated instrument for assessing BFA. Nevertheless, attempts were made to include representative questions. Lastly, as a hospital-based study, the generalizability of the findings to the broader Ugandan population may be limited although we attempted to include hospital visitors to account for this.

## Conclusions

This study reveals a concerning lack of burn first aid (BFA) knowledge and unsatisfactory practices among participants in Uganda, highlighting the need for targeted education and awareness campaigns. To improve burn care outcomes, we recommend implementing evidence-based BFA practices, regulating antibiotic use, and providing specialized training especially for acid attacks. Collaboration between policymakers, healthcare professionals, and community leaders is vital in advocacy and achieving these goals and promoting better BFA knowledge and practices.

## Data Availability

All data produced in the present study are available upon reasonable request to the authors

## Author contributions

BK was responsible for conceiving the article. BK is the guarantor. BK, JBB, FB wrote the manuscript. DK, EM,MN collected and organized the data. JBB,FB,DB and AEE critically appraised the entire project. All authors critically revised and approved the final manuscript.

## Acknowledgements

The authors would like to sincerely thank the burn patients, caregivers, and visitors for their crucial participation in this study. Special appreciation goes to Dr. Anders and Eva Nielson for their invaluable support in enabling the research on burn first aid practices in Uganda. Gratitude is also extended to plastic surgeons Dr. Kalanzi Edris and Dr. Alenyo Rose at Kiruddu National Referral Hospital for their guidance and critique. The dedicated Kiruddu Plastic Surgery Ward staff’s unwavering assistance and support throughout this research are also deeply acknowledged.

## Data sharing statement

Data available upon request.

## References

1. Kaddoura I, Abu-Sittah G, Ibrahim A, Karamanoukian R, Papazian N. Burn injury: review of pathophysiology and therapeutic modalities in major burns. Ann Burns Fire Disasters. Jun 30 2017;30(2):95–102.

2. Jeschke MG, van Baar ME, Choudhry MA, Chung KK, Gibran NS, Logsetty S. Burn injury. Nature reviews. Disease primers. Feb 13 2020;6(1):11.

3. WHO. Burns. 2018; https://www.who.int/news-room/fact-sheets/detail/burns.

4. Nthumba PM. Burns in sub-Saharan Africa: A review. Burns. Mar 2016;42(2):258–266.

5. Kobusingye O, Guwatudde D, Lett R. Injury patterns in rural and urban Uganda. Injury Prevention. 2001;7(1):46–50.

6. Asaria J, Kobusingye OC, Khingi BA, Balikuddembe R, Gomez M, Beveridge M. Acid burns from personal assault in Uganda. Burns. Feb 2004;30(1):78–81.

7. WHO. Burn prevention and Care. http://apps.who.int/iris/bitstream/handle/10665/97852/9789241596299_eng.pdf?sequenc e=1&isAllowed=y.

8. Yakupu A, Zhang J, Dong W, Song F, Dong J, Lu S. The epidemiological characteristic and trends of burns globally. BMC Public Health. 2022/08/22 2022;22(1):1596.

9. Hop MJ, Polinder S, van der Vlies CH, Middelkoop E, van Baar ME. Costs of burn care: a systematic review. Wound Repair Regen. Jul-Aug 2014;22(4):436–450.

10. Sanchez JL, Bastida JL, Martínez MM, Moreno JM, Chamorro JJ. Socio-economic cost and health-related quality of life of burn victims in Spain. Burns. Nov 2008;34(7):975–981.

11. Gerstl J, Kilgallon J, Nawabi N, et al. The Global Macroeconomic Burden of Burn Injuries. Plastic and Reconstructive Surgery – Global Open. 2021;9(10S).

12. UBPI. Uganda Burns and Plastic surgery Institure Annual report: UBPI;2014.

13. ISER. An analysis of Uganda’s Health Sector Budget2021.

14. Skinner AM, Brown TL, Peat BG, Muller MJ. Reduced hospitalisation of burns patients following a multi-media campaign that increased adequacy of first aid treatment. Burns. Feb 2004;30(1):82–85.

15. Kattan AE, Alshomer F, Alhujayri AK, Addar A, Aljerian A. Current knowledge of burn injury first aid practices and applied traditional remedies: a nationwide survey. Burns & Trauma. 2016;4(1):1–7.

16. Cuttle L, Pearn J, McMillan JR, Kimble RM. A review of first aid treatments for burn injuries. Burns. Sep 2009;35(6):768–775.

17. Galabuzi C, Agea JG, Fungo BL, Kamoga RM. Traditional medicine as an alternative form of health care system: a preliminary case study of Nangabo sub-county, central Uganda. Afr J Tradit Complement Altern Med. Oct 15 2009;7(1):11–16.

18. AlQahtani FA, Alanazi MA, Alanazi MK, Alshalhoub KS, Alfarhood AA, Ahmed SM. Knowledge and practices related to burn first aid among Majmaah community, Saudi Arabia. J Family Med Prim Care. Feb 2019;8(2):594–598.

19. Kitara D, Aloyo J, Obol J, Anywar DA. Epidemiology of burn injuries: A basis for prevention in a post-conflict, Gulu, northern Uganda: A crosssectional descriptive study design2011.

20. Arthur A. Prevalence, clinical presentation and outcome of paediatric patients with burns managed and admitted at Kiryandongo District hospitaL Uganda: a Desertation. 2018. Located at: BMS.

21. Albertyn R, Berg A, Numanoglu A, Rode H. Traditional burn care in sub-Saharan Africa: A long history with wide acceptance. Burns. 2015/03/01/ 2015;41(2):203–211.

22. Nakitto M, Lett R. Paediatric burn injuries: a hospital based study in Uganda. Injury Prevention. 2010;16(Supplement 1):A46–A47.

23. Gete BC, Mitiku TD, Wudineh BA, Endeshaw AS. Knowledge, attitude, and practice towards burn first aid and its associated factors among caregivers attending burn units in Addis Ababa, Ethiopia. A cross-sectional study. Annals of Medicine and Surgery. 2022/09/01/ 2022;81:104402.

24. Ramli RN, Prawoto A, Riasa NP, Saputro ID, Mas’Ud AF. Epidemiology and Knowledge of First Aid Treatment Related to Burn Injury in the Rural Region of Kulon Progo, Indonesia. Open Access Macedonian Journal of Medical Sciences. 2021;9(E):101–108.

25. Sonavane RS, Kasturi A, Kiran D, Kumari R. Knowledge and assessed practice regarding first aid among mothers of under 15 years children -A community based study in a rural area of south India2014.

26. Mishra SK, Mahmood S, Baig MA. Burn first aid knowledge and its determinants among general population of Rawalpindi. European Journal of Trauma and Emergency Surgery. 2019/12/01 2019;45(6):1121–1128.

27. Arifuzzaman M, Muhammad F, Farahnaz S, Chowdury A, Shahjahan M, Chowdhury A. Burn prevention and first aid knowledge among high school students in Bangladesh. Daffodil Int Univ J Allied Health Sci. 2016;3(1):41–49.

28. Lam NN, Li F, Tuan CA, Huong HTX. To evaluate first aid knowledge on burns management amongst high risk groups. Burns Open. 2017/07/01/ 2017;1(1):29–32.

29. Nakitto M, Lett R. The preparedness of Ugandan schools for fires. Injury Prevention. 2010;16(Suppl 1):A149–A149.

30. Babakir-Mina M, Othman N, Najmuldeen H, et al. Antibiotic susceptibility of vancomyin and nitrofurantoin in Staphylococcus aureus isolated from burnt patients in Sulaimaniyah, Iraqi Kurdistan. The new microbiologica. 2012;35 4:439–446.

31. Fadeyibi IO, Ibrahim NA, Mustafa IA, Ugburo AO, Adejumo AO, Buari A. Practice of first aid in burn related injuries in a developing country. Burns. Sep 2015;41(6):1322–1332.

32. Chirongoma F, Chengetanai S, Tadyanemhandu C. First aid practices, beliefs, and sources of information among caregivers regarding paediatric burn injuries in Harare, Zimbabwe: A cross-sectional study. Malawi Med J. Jun 2017;29(2):151–154.

33. Kattan AE, AlShomer F, Alhujayri AK, Addar A, Aljerian A. Current knowledge of burn injury first aid practices and applied traditional remedies: a nationwide survey. Burns Trauma. 2016;4:37.

34. Nduagubam OC, Mba UC, Onumaegbu OO, et al. Paediatric burn injuries in Enugu, South-East Nigeria: A 7-year multi-centre retrospective review. Burns. 2022/03/01/ 2022;48(2):432–439.

35. Saaiq M, Ahmad S, Zaib MS. Burn wound infections and antibiotic susceptibility patterns at pakistan institute of medical sciences, islamabad, pakistan. World J Plast Surg. Jan 2015;4(1):9–15.

